# Persistence of Pulmonary Hypertension in Patients undergoing Ventricular Assist Devices and Orthotopic Heart Transplantation

**DOI:** 10.1101/2022.08.29.22279336

**Authors:** Arun Rajaratnam, Ameen El-Swais, Charles McTiernan, Floyd W. Thoma, Moaaz O. Baghal, Kristen Raffensperger, Chung-Chou H. Chang, Gavin W. Hickey, Faraaz A. Shah, Imad Al Ghouleh

**Author notes:** **Address for Correspondence:** Imad Al Ghouleh, PhD, 1704.1 Biomedical Science Tower (Starzl BST), 200 Lothrop Street, Pittsburgh, PA 15261. **Disclosures:** All authors have no relevant financial or non-financial relationships to disclose.

## Abstract

**Background:** Pulmonary hypertension (PH) is common in advanced heart failure which exhibits short-term improvement after left ventricular assist device (VAD) implantation or orthotopic heart transplantation (OHT), but long-term effects remain unknown. This study evaluated PH persistence after VAD as destination therapy (VAD-DT), bridge to transplant (VAD-BTT), or OHT-alone.

**Methods:** A retrospective review of patients who underwent VAD-DT (n=164), VAD-BTT (n=111), or OHT-alone (n= 138) at a single tertiary-care center. Right heart catheterization (RHC) data was collected pre-, post-intervention (VAD and/or OHT), and 1-year from final intervention (latest-RHC) to evaluate the longitudinal hemodynamic course of right ventricular (RV) function and PH. PH (Group-II and Group-I) definitions were adapted from expert guidelines.

**Results:** All groups showed significant improvements in mean pulmonary artery pressure (mPAP), pulmonary artery wedge pressure (PAWP), cardiac output (CO), and pulmonary vascular resistance (PVR) at each RHC with greatest improvement at post-intervention RHC (Post-VAD or Post-OHT). PH proportion reduced from 98% to 26% in VAD-BTT, 92% to 49% VAD-DT, and 76% to 28% from pre-intervention to latest-RHC. At latest-RHC mPAP remained elevated in all groups despite normalization of PAWP and PVR. VAD-supported patients exhibited suppressed pulmonary artery pulsatility index (PaPi< 3.7) with improvement only post-transplant at latest-RHC alongside improved right atrial pressures (RAP). Furthermore, post-transplant with PH at latest-RHC (n=60) exhibited lower survival (HR: 2.1 [95%CI: 1.3-3.4], p<0.001).

**Conclusion:** Despite an overall significant improvement in pulmonary pressures and PH proportion, a notable subset of patients exhibited residual RV derangements and PH persistence post-intervention. This post-intervention PH impacted post-transplant survival.

**Condensed Abstract:** This study evaluates the longitudinal hemodynamic course of right ventricular (RV) function associated with heart-failure-related Pulmonary Hypertension (PH) and the impact of advanced interventions such as ventricular assist device and/or orthotopic heart transplant on PH persistence. Patients undergoing advanced interventions exhibit differences in PH prevalence which are dependent on the intervention received. Our results highlight despite an overall improvement in pulmonary pressures and PH, a notable number of patients exhibit RV dysfunction consistent with persistent PH. Furthermore, the persistence of PH after successful cardiac transplantation in a subset of patients continues to impact mortality in the long-term.

## Introduction

Heart failure (HF) affects more than 6 million people in the United States with a projected increase in prevalence up to 46% in the next 2 decades (1). Pulmonary hypertension (PH) is common in HF in left heart disease (PH-LHD, Group II PH) with a reported prevalence ranging from 54% to 83% in HF with preserved ejection fraction (HFpEF) (2, 3) and 73% in HF with reduced ejection fraction (HFrEF)(4). Previous studies have reported an increased 30-day post-transplant mortality associated with right ventricular (RV) failure in Chronic HF patients with pre-transplant PH(5, 6). Patients with end-stage HF who develop PH exhibit refractory symptoms despite optimized medical therapy and may require specialized intervention such as orthotopic heart transplantation (OHT) or mechanical circulatory devices (MCD). Despite notable improvements in the prevention and treatment of HF-related risk factors and tailored medical therapy (1, 7), PH-LHD patients still comprise a stratum with high risk of mortality and hospitalization (2, 8–10).

Although rates of heart transplantation have steadily increased over the last decade, the number of patients with advanced heart failure (New York Heart Classification 3a-V,NYHA) remains significantly greater than the number of available donor hearts (11); subsequently, ventricular assist devices (VADs) emerged as an important therapeutic modality in advanced HF with > 20,000 implantations over the last 2 decades (12). VADs are MCDs that augment left ventricular (LV) function by improving cardiac output in patients with HFrEF and are used as destination therapy (DT) or bridge to transplantation (BTT) in patients evaluated for OHT. VADs improve RV and pulmonary pressures in the short and intermediate term (30–180 days) in PH-LHD, however, studies with longitudinal hemodynamic data are small, with a primary focus on reversal of elevated PVR given increased likelihood of post-operative RV failure (13–15). VADs have improved survival in chronic HF (12, 16), however, RV failure still negatively impacts survival in VAD-assisted patients(17). Interestingly, BTT patients experienced decreased survival at 1-year post-transplant associated with higher mean pulmonary artery pressure (mPAP) and pulmonary vascular resistance (PVR) compared to OHT-alone (18). Therefore, patients managed with VADs may exhibit chronic PH-LHD albeit with lower RV and pulmonary pressures, which may plausibly contribute to higher morbidity and mortality. The paucity of long-term hemodynamic data with respect to PH-LHD in patients post-VAD or OHT limits evaluation of RV adaptation to chronic pressure overload (i.e HF) required to characterize PH persistence. Thus, we evaluated the prevalence and the natural hemodynamic course of PH and RV function in patients eligible for OHT and VAD to understand the potential impact of PH in patients with advanced HF.

## Methods

This study is a retrospective analysis of 413 patients with advanced HF who underwent evaluation and received intervention from 2008 – 2021 for OHT or VAD at University of Pittsburgh Medical Center (UPMC). Designation of BTT or DT was determined based on review of medical records of treatment received by the patient by two independent reviewers (A.R and A.E-S). This study was approved by the Institutional Review Board at the University of Pittsburgh (STUDY20090170). Baseline demographics, comorbidities, and laboratory data were collected within 1 month of first right heart catheterization (RHC) prior to initial intervention (“pre-intervention”) defined as RHC prior to VAD or OHT. Hemodynamic data from RHC and echocardiography were obtained within a median of 30±7 days prior to intervention, 3±1 months post-intervention (post-VAD in VAD-DT or post-OHT in OHT alone, and after each intervention in BTT), and 12±4 months after last intervention (latest-RHC in VAD or OHT).

### Evaluation of Invasive Hemodynamics

HF was categorized by LV ejection fraction (LVEF) with HFrEF defined as EF≤40%. PH definitions were adapted from expert guidelines (19) and derived from RHCs at rest in the supine position (Suppl. Figure 4). Pre-capillary PH (Pulmonary Arterial Hypertension, PAH or Group I PH) was defined as mPAP>20 mmHg, pulmonary artery wedge pressure (PAWP) < 15mmHg & PVR≥3 Woods Unit (WU). PH-LHD (Group II PH) was defined as mPAP > 20 mmHg & PAWP ≥15mmHg and further sub-categorized into Isolated post-capillary PH (IpcPH) defined by PVR <3 WU and Combined post-capillary PH (CpcPH) defined as PVR>3 WU. Isolated elevated mPAP was defined as mPAP≥20 mmHg, PAWP < 15mmHg and PVR < 3 WU, and normal hemodynamics (i.e., No PH) defined as mPAP < 20mmg, PAWP < 15mmHg and PVR < 3 WU. PH was defined as PAH & PH-LHD emphasizing total PH burden irrespective of pathophysiology. RHC measures including mPAP, PAWP, right atrial pressure (RAP), cardiac output (CO, by thermodilution), pulmonary artery systolic pressure (PASP), pulmonary artery diastolic pressure (PADP), and heart rate (HR) were obtained to calculate Pulmonary Artery Pulsatility Index (PAPi, calculated as PASP-PADP/Stroke Volume (SV)), Pulmonary artery elastance (Pae, calculated as PASP-PAWP/SV), Pulmonary artery compliance (PAC, calculated as SV/ Pulmonary Artery Pulse Pressure, the latter defined as PASP-PADP), and PVR (calculated as mPAP-PAWP/CO).

### Statistical analyses

Data are expressed as mean with standard deviation (SD) for normally distributed continuous variables or median with interquartile range (IQR) for skewed continuous variables, and absolute value with percent for categorical variables. Comparison of variables between treatment groups used Students-t test or One-way analysis of variance (with Welch’s t-test) for continuous variables, Mann-Whitney U test or Kruskal-Wallis test for skewed continuous variables, and Chi-square test or Fisher’s exact test for categorical variables where appropriate. Generalized linear models using the generalized estimating equation (GEE) was utilized to evaluate differences in repeated measures of hemodynamic variables and proportion of PH within each treatment group and utilized the Wald test to evaluate statistical significance. GEE with auto-regression working correlation matrices was used to reflect the correlation among the outcomes over the different time periods (RHC in longitudinal order). Comparisons among time periods (RHC order) were conducted using the Bonferroni correction for multiplicity. Complete cases were used for the primary analysis. Imputation was considered but not pursued owing to the longitudinal focus of our study and avoiding overestimation of hemodynamic values and proportions similar to previous studies (20). Missing data post-VAD in VAD-DT was predominantly attributable to mortality. Missing data post-VAD in VAD-BTT was considered to be random based on distribution of availability of RHC data. Kaplan-Meier method was used to estimate unadjusted survival curves for each treatment group and the log-rank test was used to compare among survival curves. Cox proportional-hazards models were used to estimate hazard ratio (HR) and 95% confidence intervals (CI) between groups. Proportional hazards assumption was assessed using Schoenfeld residual plots. Data management and analysis were performed using SPSS Version 27.0 (IBM Corp., Armonk NY USA) and R statistical package (version 4.0.2, R Core Team, 2022). Figures and illustrations were produced using Prism 9 (GraphPad Software, San Diego, California USA), Biorender (Biorender.com), and the R statistical package. P-value <0.05 was considered statistically significant.

## Results

A total of 413 patients were evaluated for VAD and OHT during the study period: 111 patients underwent VAD-BTT, 164 VAD-DT, and 138 had OHT-alone. Overall, patients were predominantly male (78%), Caucasian (81%) with no significant differences in co-morbidities (Table 1 and Suppl. Table 1), and all were classified as NYHA class IIIb-IV. Significant differences in age were observed in VAD-BTT patients as they received VAD and OHT relatively younger than VAD-DT or OHT-alone patients. OHT-alone patients had the lowest BMI relative to VAD-DT and VAD-BTT (Table 1).

**Table 1.**
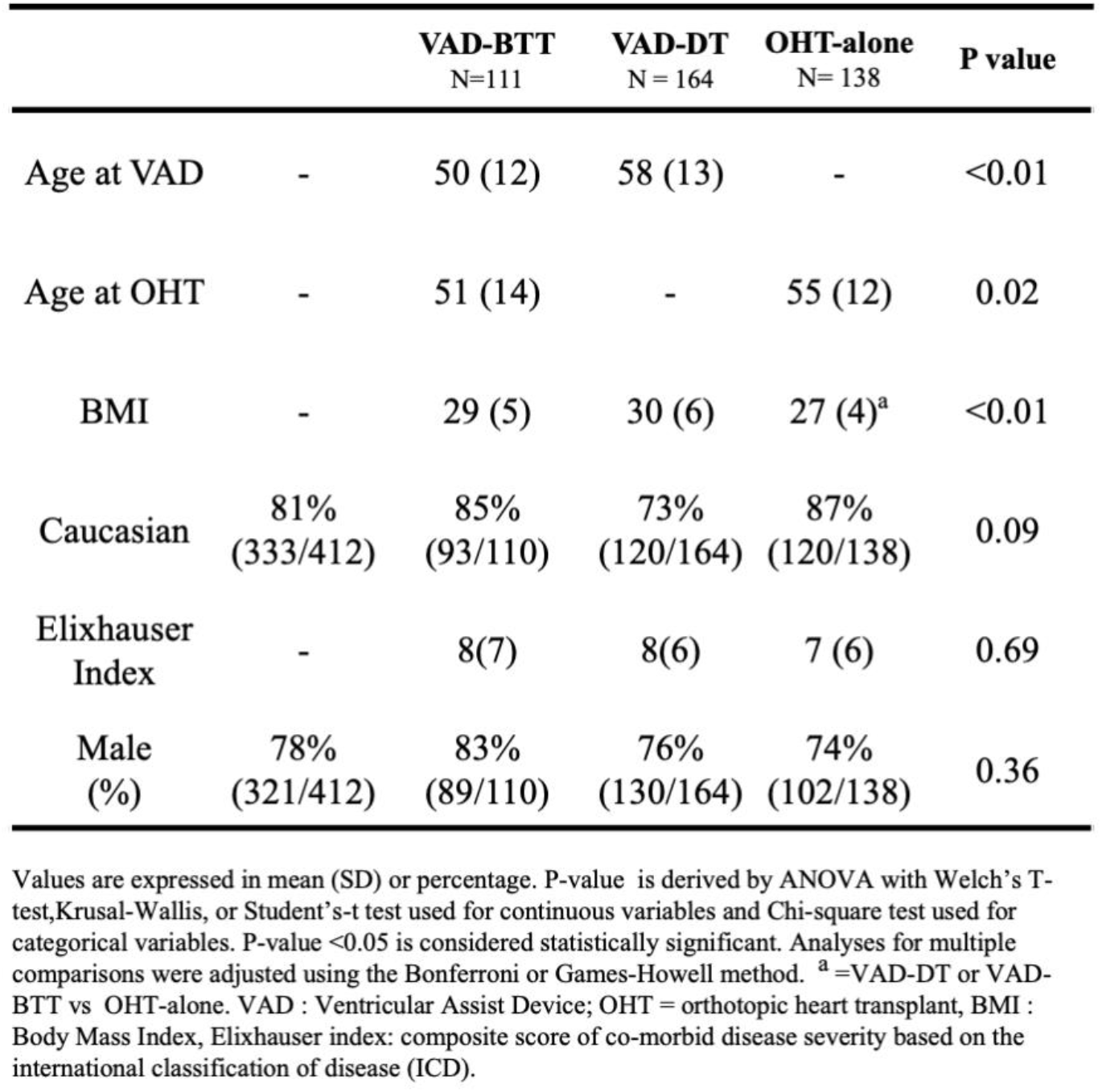
Baseline Demographics.

|  |  | **VAD-BTT**<br>N=111 | **VAD-DT**<br>N = 164 | **OHT-alone**<br>N= 138 | **P value** |
| --- | --- | --- | --- | --- | --- |
| Age at VAD | - | 50 (12) | 58 (13) | - | <0.01 |
| Age at OHT | - | 51 (14) | - | 55 (12) | 0.02 |
| BMI | - | 29 (5) | 30 (6) | 27 (4)<sup>a</sup> | <0.01 |
| Caucasian | 81%<br>(333/412) | 85%<br>(93/110) | 73%<br>(120/164) | 87%<br>(120/138) | 0.09 |
| Elixhauser Index | - | 8(7) | 8(6) | 7 (6) | 0.69 |
| Male (%) | 78%<br>(321/412) | 83%<br>(89/110) | 76%<br>(130/164) | 74%<br>(102/138) | 0.36 |

Values are expressed in mean (SD) or percentage. P-value is derived by ANOVA with Welch's T-test, Kruskal-Wallis, or Student's-t test used for continuous variables and Chi-square test used for categorical variables. P-value <0.05 is considered statistically significant. Analyses for multiple comparisons were adjusted using the Bonferroni or Games-Howell method. <sup>a</sup> =VAD-DT or VAD-BTT vs OHT-alone. VAD : Ventricular Assist Device; OHT = orthotopic heart transplant, BMI : Body Mass Index, Elixhauser index: composite score of co-morbid disease severity based on the international classification of disease (ICD).

Presence of PH at pre-intervention RHC was high in all groups: 98% in VAD-BTT, 92% in VAD-DT, and 76% in OHT-alone (Figure 1). At pre-intervention, presence of PAH was < 5% in all treatment groups (Supp Fig 1A-C). Independent of treatment received, there was an overall significant reduction in the proportion of PH during the period of study (Figure 1). VAD-BTT patients exhibited reduction of PH from 98% at pre-intervention to 48% at post-VAD RHC; however, at post-OHT and latest-RHC, a trend towards further reduction in PH was observed but was not statistically significant (Figure 1).PH in VAD-DT improved to 68% at post-VAD with a non-significant reduction to 49% at latest-RHC. OHT-alone patients showed significant reduction in PH at each RHC and 28% had PH at latest-RHC. The predominant hemodynamic sub-phenotypes of PH at pre-intervention in VAD-BTT were equally IpcPH and CpCH at 48% and 46%, respectively (Suppl. Figure 1A), with similar observations in VAD-DT (44% with IpcPH and CpcPH, Suppl. Figure 1B). However, in OHT-alone, 50% of patients had IpcPH while 22% had CpcPH (Suppl. Fig 1C). At latest-RHC, IpcPH was present at 19% in BTT and 17% in OHT-alone with no CpcPH in BTT and 1% in OHT-alone (Suppl.Figure 1A,C). VAD-DT exhibited the highest proportion of patients with persisting IpcPH and CpcPH at 33% and 18%, respectively (Suppl. Figure1B).

**Figure 1.**
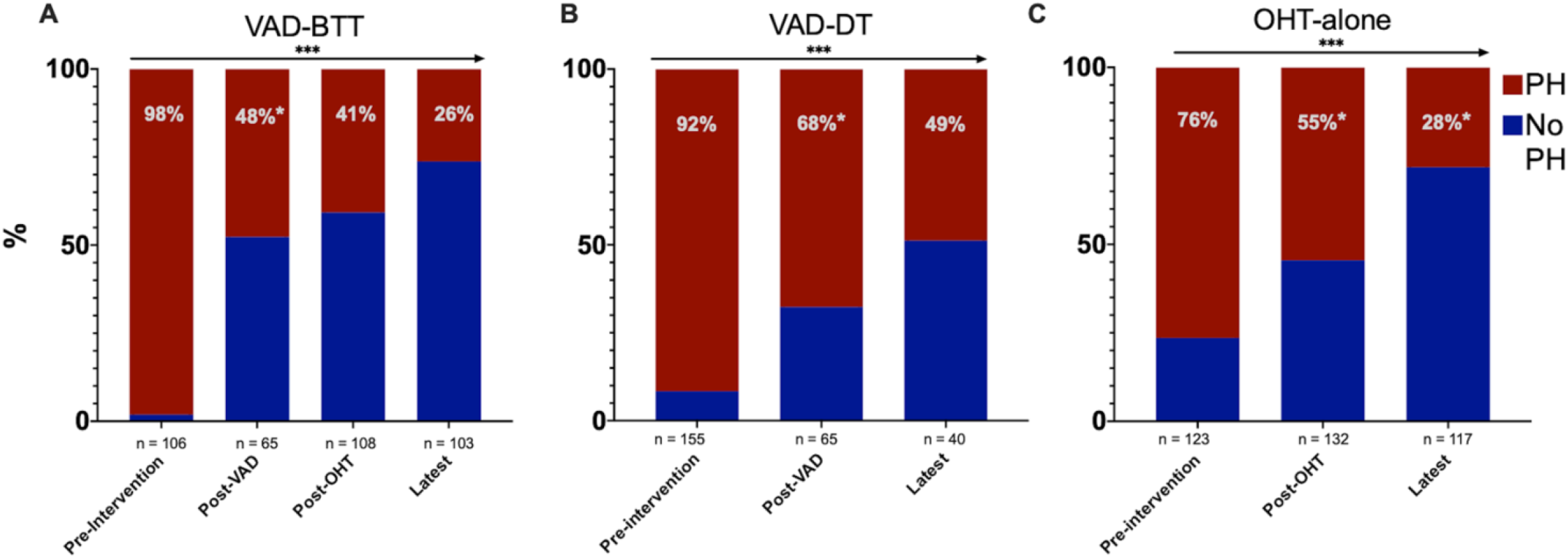
Prevalence of Pulmonary Hypertension on Right Heart Catheterization on Pre- and Post-Intervention VAD as Bridge to Transplant (VAD-BTT,panel A), Destination Therapy (VAD-DT,panel B), and Orthotopic transplant only (OHT-alone, panel C). Panel arrow denotes linear reduction in the proportion of pulmonary hypertension (PH) at successive RHC by treatment group with statistical significance < 0.001 (***) by the generalized estimating equation utilizing the Wald test. * denotes within group comparison(current bar compared to previous bar) by the Bonferroni method. P values <0.05 was considered statistically significant. PH = Pulmonary Hypertension.

Hemodynamic evaluation of RV function throughout the study showed significant reduction in all measured variables in all groups with exception of PAPi in VAD-DT (Table 2). Patients who underwent VAD-BTT had significant reductions in pulmonary pressures overall but exhibited abnormal mPAP, PAWP, RAP, and PaPi related to treatment received at specific RHCs (Figure 2A, Table 2). Elevated mPAP and RAP were observed despite PVR < 3 WU and normal PAWP at post-VAD and post-OHT in VAD-BTT. The non-significant improvement in mPAP, PAWP, and RAP at post-OHT despite successive improvement in PVR was surprising, but is likely related to reduced CO suggestive of diminished RV recovery in the post-operative period (Table 2). Significant improvement in mPAP, RAP, and PAWP were observed at latest-RHC, and specifically in PAPi which was suppressed (<3.7) at previous RHCs (Table 2). PAe and PAC significantly improved post-VAD and PAe latest-RHC independent of PVR. However, at latest-RHC a notable proportion of patients exhibited elevated mPAP and RAP with suppressed PaPi (Figure 2A and Table 3).

**Table 2.**
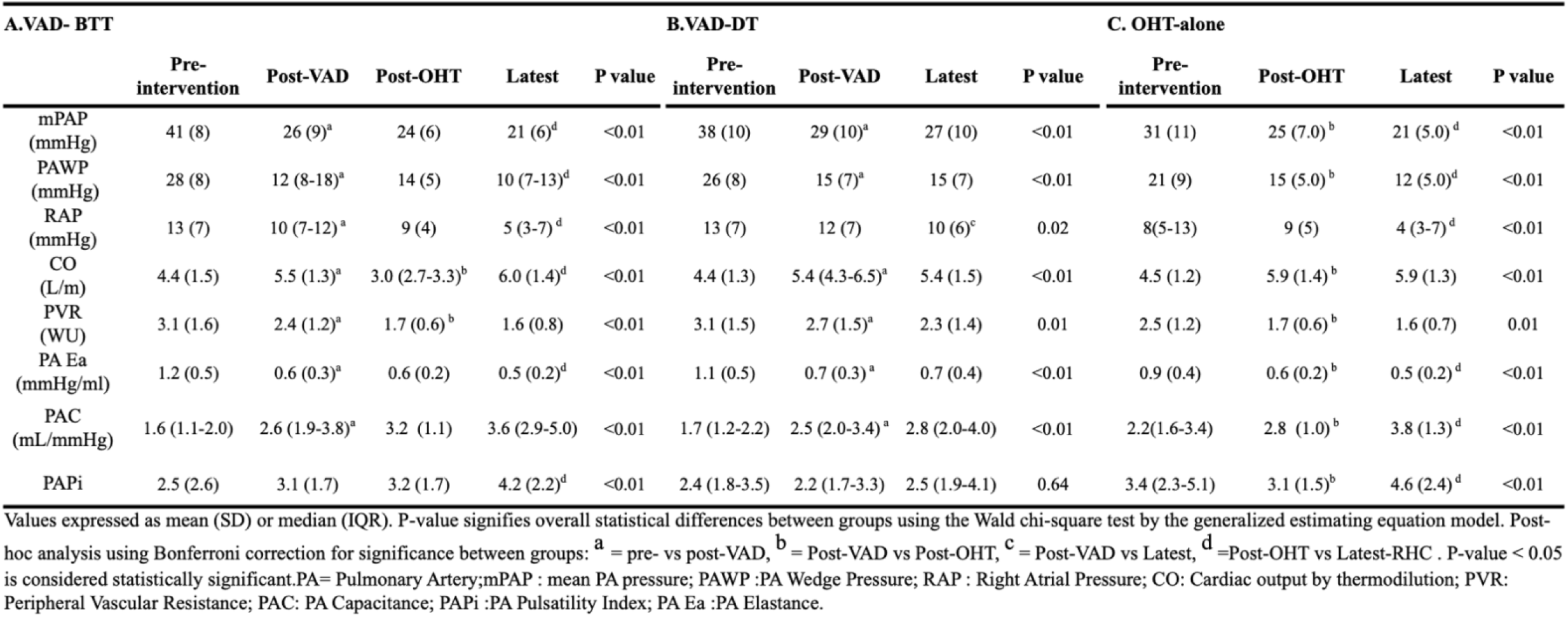
Longitudinal Right Heart Catheterization-based Hemodynamics by Treatment Groups.

| A.VAD- BTT |  |  |  |  |  | B.VAD-DT |  |  |  | C. OHT-alone |  |  |  |
| --- | --- | --- | --- | --- | --- | --- | --- | --- | --- | --- | --- | --- | --- |
|  | Pre-intervention | Post-VAD | Post-OHT | Latest | P value | Pre-intervention | Post-VAD | Latest | P value | Pre-intervention | Post-OHT | Latest | P value |
| mPAP (mmHg) | 41 (8) | 26 (9)<sup>a</sup> | 24 (6) | 21 (6)<sup>d</sup> | <0.01 | 38 (10) | 29 (10)<sup>a</sup> | 27 (10) | <0.01 | 31 (11) | 25 (7.0)<sup>b</sup> | 21 (5.0)<sup>d</sup> | <0.01 |
| PAWP (mmHg) | 28 (8) | 12 (8-18)<sup>a</sup> | 14 (5) | 10 (7-13)<sup>d</sup> | <0.01 | 26 (8) | 15 (7)<sup>a</sup> | 15 (7) | <0.01 | 21 (9) | 15 (5.0)<sup>b</sup> | 12 (5.0)<sup>d</sup> | <0.01 |
| RAP (mmHg) | 13 (7) | 10 (7-12)<sup>a</sup> | 9 (4) | 5 (3-7)<sup>d</sup> | <0.01 | 13 (7) | 12 (7) | 10 (6)<sup>c</sup> | 0.02 | 8(5-13) | 9 (5) | 4 (3-7)<sup>d</sup> | <0.01 |
| CO (L/m) | 4.4 (1.5) | 5.5 (1.3)<sup>a</sup> | 3.0 (2.7-3.3)<sup>b</sup> | 6.0 (1.4)<sup>d</sup> | <0.01 | 4.4 (1.3) | 5.4 (4.3-6.5)<sup>a</sup> | 5.4 (1.5) | <0.01 | 4.5 (1.2) | 5.9 (1.4)<sup>b</sup> | 5.9 (1.3) | <0.01 |
| PVR (WU) | 3.1 (1.6) | 2.4 (1.2)<sup>a</sup> | 1.7 (0.6)<sup>b</sup> | 1.6 (0.8) | <0.01 | 3.1 (1.5) | 2.7 (1.5)<sup>a</sup> | 2.3 (1.4) | 0.01 | 2.5 (1.2) | 1.7 (0.6)<sup>b</sup> | 1.6 (0.7) | 0.01 |
| PA Ea (mmHg/ml) | 1.2 (0.5) | 0.6 (0.3)<sup>a</sup> | 0.6 (0.2) | 0.5 (0.2)<sup>d</sup> | <0.01 | 1.1 (0.5) | 0.7 (0.3)<sup>a</sup> | 0.7 (0.4) | <0.01 | 0.9 (0.4) | 0.6 (0.2)<sup>b</sup> | 0.5 (0.2)<sup>d</sup> | <0.01 |
| PAC (mL/mmHg) | 1.6 (1.1-2.0) | 2.6 (1.9-3.8)<sup>a</sup> | 3.2 (1.1) | 3.6 (2.9-5.0) | <0.01 | 1.7 (1.2-2.2) | 2.5 (2.0-3.4)<sup>a</sup> | 2.8 (2.0-4.0) | <0.01 | 2.2(1.6-3.4) | 2.8 (1.0)<sup>b</sup> | 3.8 (1.3)<sup>d</sup> | <0.01 |
| PAPi | 2.5 (2.6) | 3.1 (1.7) | 3.2 (1.7) | 4.2 (2.2)<sup>d</sup> | <0.01 | 2.4 (1.8-3.5) | 2.2 (1.7-3.3) | 2.5 (1.9-4.1) | 0.64 | 3.4 (2.3-5.1) | 3.1 (1.5)<sup>b</sup> | 4.6 (2.4)<sup>d</sup> | <0.01 |

Values expressed as mean (SD) or median (IQR). P-value signifies overall statistical differences between groups using the Wald chi-square test by the generalized estimating equation model. Post-hoc analysis using Bonferroni correction for significance between groups: <sup>a</sup> = pre- vs post-VAD, <sup>b</sup> = Post-VAD vs Post-OHT, <sup>c</sup> = Post-VAD vs Latest, <sup>d</sup> =Post-OHT vs Latest-RHC . P-value < 0.05 is considered statistically significant.PA= Pulmonary Artery;mPAP : mean PA pressure; PAWP :PA Wedge Pressure; RAP : Right Atrial Pressure; CO: Cardiac output by thermodilution; PVR: Peripheral Vascular Resistance; PAC: PA Capacitance; PAPi :PA Pulsatility Index; PA Ea :PA Elastance.

**Table 3.** Hemodynamics at latest-RHC with proportion of patients with abnormal measures.

|  | VAD-BTT<br>(n = 101) | VAD-DT<br>(n= 41) | OHT-alone<br>(n =115) | P value |
| --- | --- | --- | --- | --- |
| mPAP<br>(mmHg) | 48% | 68%<sup>a</sup> | 49% | 0.06 |
| PAWP<br>(mmHg) | 19% | 49%<sup>a</sup> | 19% | <0.01 |
| RAP<br>(mmHg) | 39% | 71%<sup>a</sup> | 27% | <0.01 |
| PAPi | 45% | 65%<sup>a</sup> | 39% | 0.02 |
| PVR<br>(WU) | 6% | 29%<sup>a</sup> | 4% | <0.01 |

**Panel A: Percent of patients with abnormalities in key hemodynamic variables at latest-RHC across treatment groups.** Values are reported as percentage of patients above or below cut-off for mPAP > 20mmHg, PAWP >15mmHg, PVR < 3WU, and PaPi < 3.7. P-value is derived using Chi-square or Fisher's exact test. P-value < 0.05 was considered significant. Analyses for multiple comparisons were adjusted using the Bonferroni method. <sup>a</sup> = denotes VAD-BTT or OHT-alone vs VAD-DT.

**Figure 2A:**
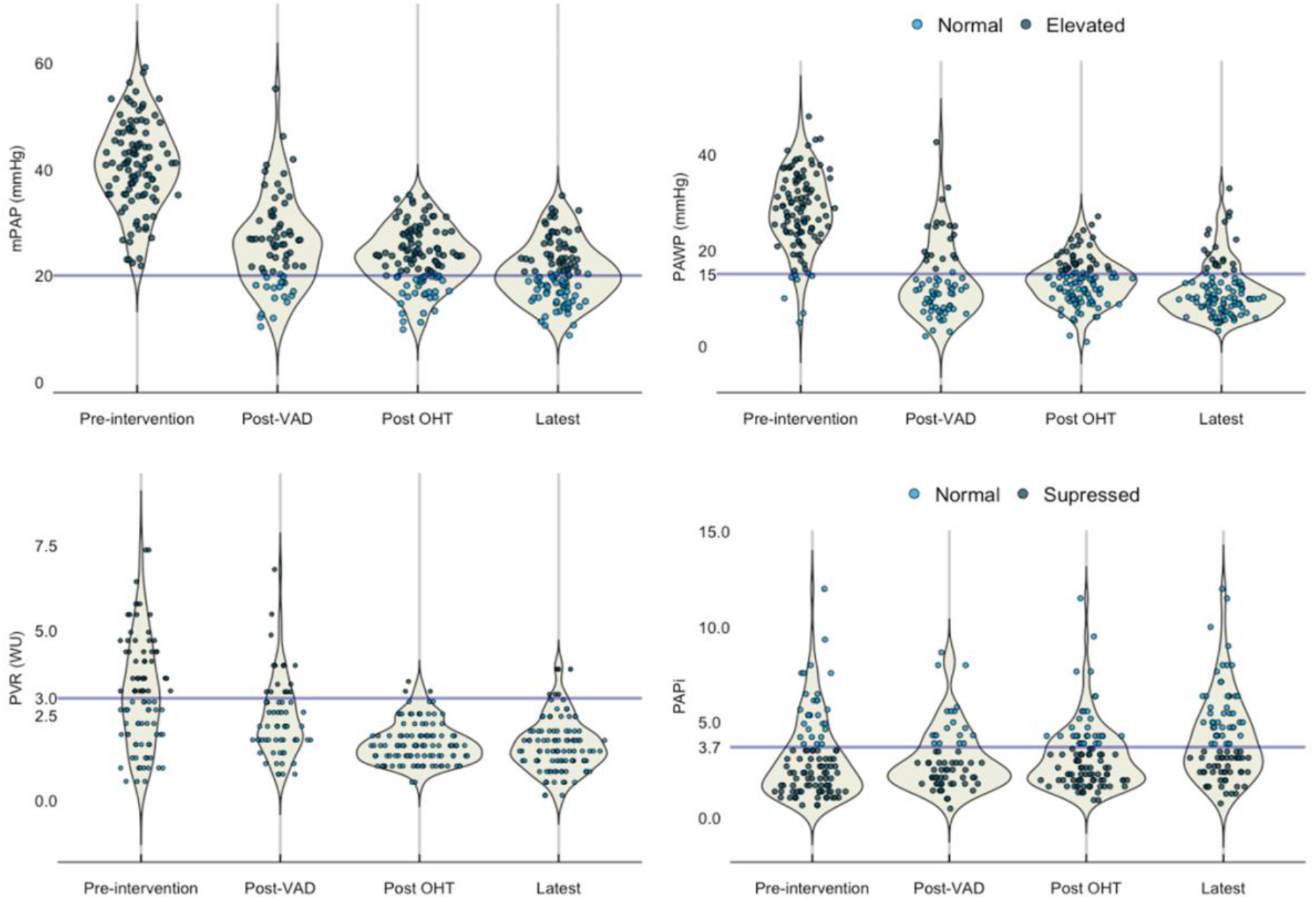
Distribution and proportion of patients with normal vs. abnormal measures in selected hemodynamic variables in the BTT group. Consistent reduction in key hemodynamic measures were observed during the study period with a notable number of patients with abnormal values at latest-RHC (abnormal mPAP and PaPi despite improved PAWP and PVR). Blue line signifies cut-off values delineating normal vs. abnormal or normal vs. suppressed in PaPi. mPAP : mean pulmonary artery pressure; PAPi :Pulmonary Artery Pulsatility Index; PAWP :Pulmonary Artery Wedge Pressure; PVR: Peripheral Vascular Resistance.

**Figure 2B:**
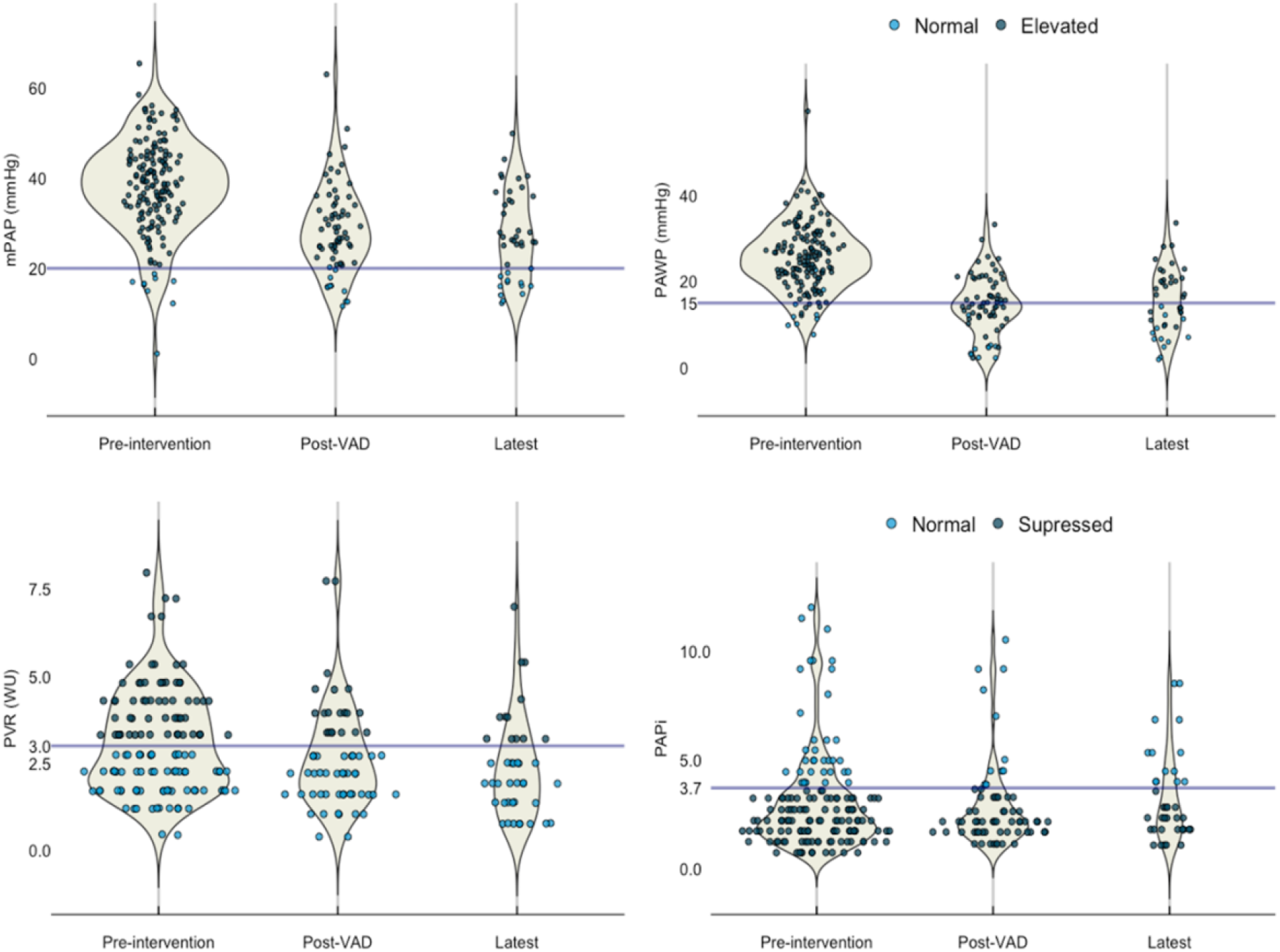
Distribution and proportion of patients with normal vs. abnormal measures in selected hemodynamic variables in the DT group. Consistent reduction in key hemodynamic measures were observed during the study period with a notable number of patients with abnormal values at latest-RHC (abnormal mPAP and PaPi despite improved PAWP and PVR). Blue line signifies cut-off values delineating normal vs. abnormal or normal vs. suppressed in PaPi. mPAP : mean pulmonary artery pressure; PAPi :Pulmonary Artery Pulsatility Index; PAWP :Pulmonary Artery Wedge Pressure; PVR: Peripheral Vascular Resistance.

**Figure 2C:**
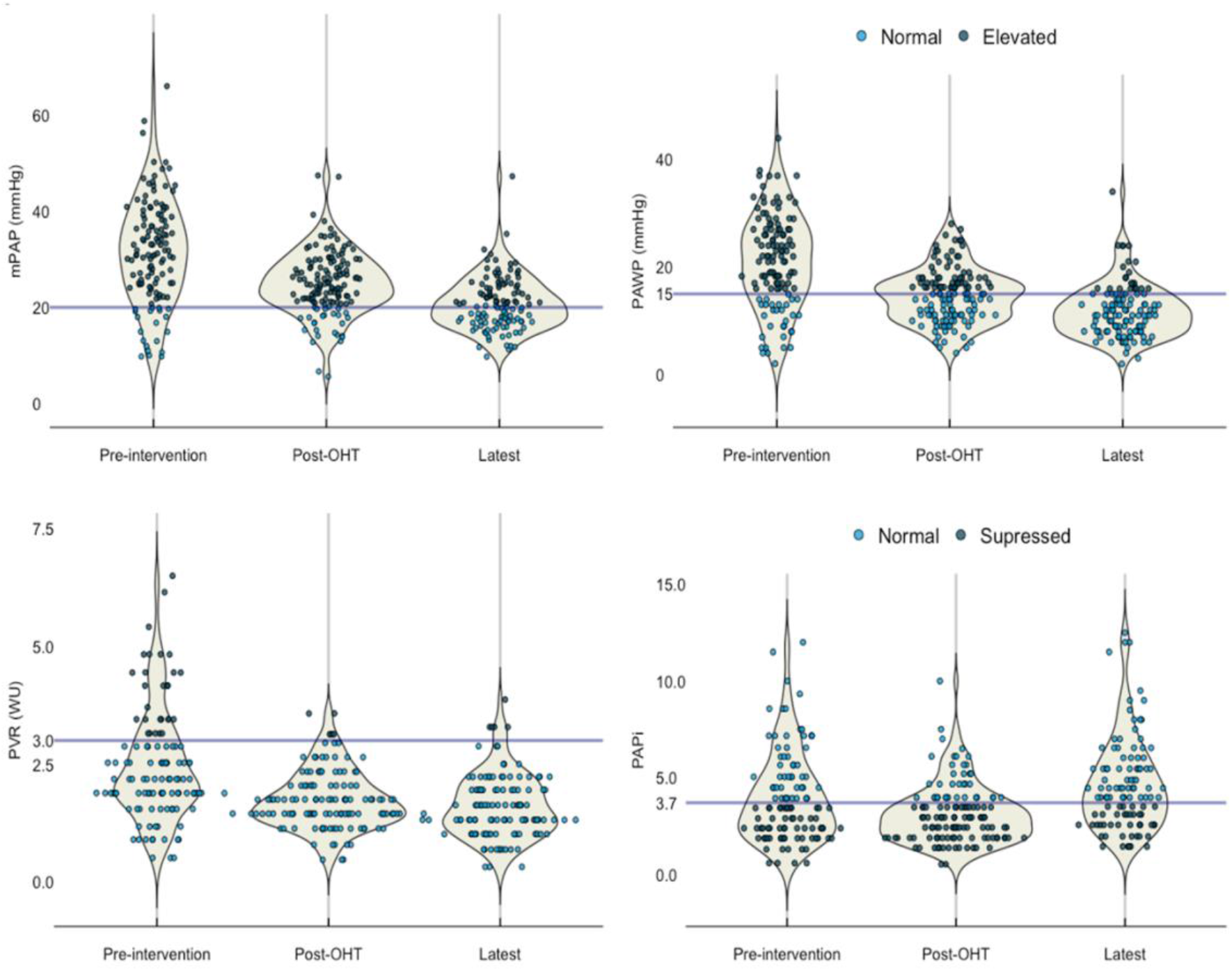
Distribution and proportion of patients with normal vs. abnormal measures in selected hemodynamic variables in the OHT group. Consistent reduction in key hemodynamic measures were observed during the study period with a notable number of patients with abnormal values at latest-RHC (abnormal mPAP and PaPi despite improved PAWP and PVR). Blue line signifies cut-off values delineating normal vs. abnormal or normal vs. suppressed in PaPi. mPAP: mean pulmonary artery pressure; PAPi :Pulmonary Artery Pulsatility Index; PAWP :Pulmonary Artery Wedge Pressure; PVR: Peripheral Vascular Resistance.

Despite improvements, patients undergoing VAD-DT had elevated mPAP with borderline elevation of PAWP post-VAD despite reduction of PVR (Table 2). No additional reductions in pulmonary pressures were noted at latest-RHC except RAP, which nonetheless remained abnormally elevated. PaPi in the DT group did not improve throughout the study despite significant improvements in PAe and PAC post-VAD which remained unchanged till latest-RHC; this is further highlighted by the distribution and proportion of abnormal values of key hemodynamic variables at each RHC in VAD-DT (Figure 2B). At latest-RHC, VAD-DT exhibited a large proportion of abnormal mPAP, PAWP, RAP, and PAPi (Figure 2B and Table 3).

Patients undergoing OHT-alone had the lowest overall pre-intervention pulmonary pressures and RV-specific parameters (Table 2). This group had continued improvement in all measures at successive RHC except RAP which only normalized in a majority of patients at latest-RHC. Like VAD-BTT, post-OHT mPAP surprisingly remained slightly elevated with borderline elevated PAWP despite improved PVR. Notably, the OHT-alone group had the lowest PVR overall at pre-intervention. PaPi was reduced post-OHT even with improved CO and is likely secondary to post-surgical effects on the RV as RAP remained unchanged as well until latest-RHC. Distribution of key hemodynamic variables at each RHC exhibited a notable proportion of patients with abnormal values despite an improvement in averaged pressures (Fig 2C and Table 3). At latest-RHC, mPAP was elevated in 49% of patients and reduced PAPi persisted in 39% similar to 45% in VAD-BTT (Table 3). PAe and PAC showed continued improvement at each RHC in the setting of PVR < 3 WU (Table 2).

At 1-year post-OHT, a total of 60 patients exhibited PH on RHC. These patients had lower survival compared to patients with no hemodynamically defined PH (No PH; Figure 3, HR: 2.1 [95% CI: 1.3-3.4], p<0.001). Median survival probability was 96% (CI:92-99%) in the No PH group and 90% (83-98%) in the PH group at 1-year and was 69% (61-78%) and 46% (33-62%) respectively at 10-years from latest-RHC. We did not observe early mortality in patients exhibiting PAH at latest-RHC during the study period (suppl. Figure 2), limiting the possibility of an effect of PAH-associated early mortality on survival in this cohort. BMI remained a significant risk factor of survival in Cox proportional hazard modeling (p<0.01), but age or previous VAD-implantation were not. We observed a reduction in survival associated with higher PAWP (≥15mmHg, Fig 4) and in patients with PH-LHD (PAH excluded), who exhibited reduction in median survival (7.5 years) with a HR of 2.2 (95% CI: 1.3-3.7, P: 0.002).

**Figure 3:**
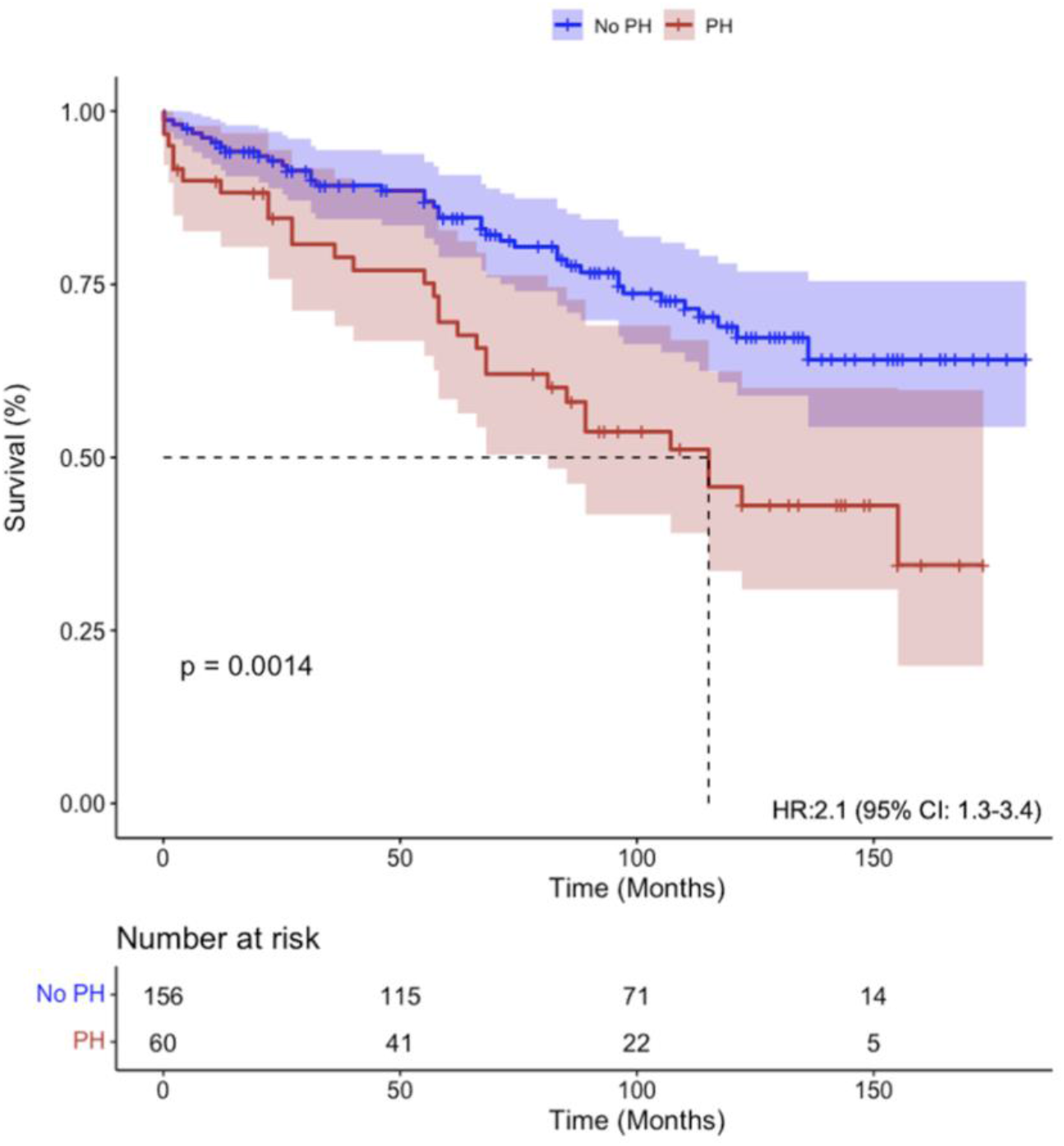
Kaplan-Meier curves in post-transplant patients with and without PH. Unadjusted survival analysis of patients with PH (Total PH) versus No PH shows significant differences in survival over the 14-year study period. Hazard ratio (HR) is derived using the Cox Proportional Hazards method with P-value using the log-rank test.

## Discussion

In a cohort of patients with advanced HF who underwent VAD, OHT, or both we demonstrate persistence of PH despite longitudinal improvements in hemodynamics and identified IpcPH as the predominant subtype in all groups. Additionally, we observe BTT patients show additive improvements in pulmonary pressures after OHT and complete amelioration of CpcPH in all post-OHT patients. We observe that despite expected normalization of LVEF (Suppl. Fig 3) and reduced PAWP at latest-RHC in most post-OHT patients, a significant proportion still had abnormal mPAP, RAP, and PAPi (Fig. 2, & Table 3). Finally, we observe PH persistence in a subset of patients at 1-year post-transplant (from both VAD-BTT and OHT-alone cohorts) was associated with an increased risk of mortality with 50% of patients surviving at 9.5-years compared to 64% without post-transplant PH.

The RV is a thin walled, crescent shaped, and low-pressure conduit system which is embryologically (21) and physiologically different from the LV(22). The RVs adaptation to pressure and volume remains tightly coupled to the pulmonary vasculature to maintain optimal cardiac function in normal and disease states, including PH(23), requiring RHC data to diagnose PH and evaluate RV function. PH persistence after advanced intervention (VAD or OHT) in end-stage HF remains an important area of study given its impact on adaptive and maladaptive RV responses which affect survival and morbidity(24). However, heterogeneity in PH definitions used (14, 19, 25), updated expert definitions of PH-LHD, and variable sample sizes with longitudinal hemodynamic data has limited the evaluation of PH persistence. Within our entire cohort, at pre-intervention CpcPH and IpcPH was prevalent in 37% and 47% of all patients, respectively; by latest-RHC, IpcPH persisted in 17-33% of all patients dependent on the treatment group, whereas CpcPH comparatively resolved in post-OHT patients (CpcPH reduced to 18% in VAD-DT patients). Resolution of CpcPH post-OHT in VAD-BTT and OHT-alone groups was likely secondary to selection of patients with PVRs < 5WU to reduce likelihood of post-operative early and late RV failure (11, 26). Patients who underwent VAD-DT were near equivalent to VAD-BTT in terms of pulmonary hemodynamics at pre-intervention and likewise showed reduced PH burden cumulatively albeit with higher proportions of IpcPH and CpcPH at latest-RHC; residual PH in VAD-DT may potentially be related to duration of PH-LHD (advanced age) and confounding with VAD-related mortality (120/164 by latest-RHC, Suppl. Table 3). The proportion of IpcPH as the predominant phenotype was unexpected but may suggest underlying chronic RV remodeling in HF without pulmonary remodeling (PVR < 3). The mechanisms leading to the development of CpcPH versus IpcPH in PH-LHD remain elusive but are hemodynamically delineated by degree of pulmonary vascular remodeling (PVR).Previous studies have shown CpcPH exhibiting histopathological and genetic polymorphisms resembling PAH, with IpcPH sharing similar vascular morphology without significant intimal or adventitial fibrosis, vessel occlusion, or plexiform vasculopathy pathognomonic for PAH (25, 27). Therefore, CpcPH and IpcPH may represent distinct parallel pathologies co-opting RV and pulmonary vascular adaptive mechanisms triggered by passive, elevated backpropagating cardiac filling pressures as the antecedent etiology; however, VAD-related LV unloading of elevated cardiac filling pressures may have a favorable reversal of pulmonary vascular remodeling with a limited, indirect impact on RV adaptation in HF(28) leading to a greater relative reduction CpcPH compared to IpcpH. Reversal of CpcPH or IpcPH are likely highly dependent on genetic predisposition and intervention(s) received in addition to mechanisms related to PH development as described above, which highlights the need for tailored, patient-specific surveillance for RVD especially in the post-intervention setting (VAD and/or OHT). Whether LV unloading by VAD reverses pulmonary vascular remodeling, increases vascular tone, or both is beyond the scope of this study, but are potential mechanisms impacting PH persistence and are areas of future research. The observed reduction of CpcPH could also be confounded by careful candidate selectivity for OHT along with potentially increased pharmacological interventions (e.g vasodilators) to reduce PVR. Though focus on CpcPH is an important consideration for VAD or OHT given its implications on mortality, we observe the IpcPH subtype to be insidious with a similar impact on patients with advanced HF.

RV function is sensitive to changes in afterload in acute and chronic pressure overload when compared to LV in HF (28). Therefore, any improvement in vascular resistance in the pulmonary circulation by VAD should translate to improved RV function. In our cohort, post-VAD residual RVD is characterized by increased mPAP, RAP, and suppressed PaPi (<3.7)(29) despite effective VAD-related unloading. PaPi is a surrogate measure of RV CO developed for VAD patients given VAD mechanically augments LV function(30). In our cohort, PaPi does not change during VAD support, suggestive of residual RVD, and significant improvement (> 4.0) is only observed 1-year after transplant (latest-RHC). Furthermore, patients with elevated mPAP post-intervention may be at continued, increased risk of mortality, morbidity, and progression of PH likened to studies evaluating patients with ‘borderline’ or ‘high-risk’ PH(31). Effective LV unloading is further demonstrated by improvements in PAe and PAC which are corollary measures of RV afterload alongside PVR which hemodynamically describe the dynamic (pulsatile) and static (resistive) forces of pulmonary resistance which independently impact survival in PH (32, 33). Altogether, we observe a significant improvement consistent with reversal of the pulmonary vascular adaptation independent of intervention. Reductions in mPAP, PAWP, and RAP at post-VAD & OHT are observed, but these measures have not altogether normalized in a notable proportion of patients (Table 3) suggestive of residual RVD especially in the latest-RHC setting; Together, these findings highlight the limited reversal of RV adaptation consistent with persistent PH observed within the entire cohort and previously in post-VAD patients. (28, 34).

Patients having undergone OHT-alone exhibited lower burden of PH at pre- and latest-RHC and showed notable improvements in successive RHCs. Nonetheless, at latest-RHC in both OHT-alone and VAD-BTT groups, despite improved pulmonary pressures 1-year post-cardiac transplant, a notable proportion of patients had abnormal RHC measures (Figure 2 and Table 3) with an average of 26-28% with persistent PH; these patients had a two-fold increase in mortality over a 14-year study period. This translated to a 9.5-year median survival time in patients with PH post-OHT where 50% survived relative to a 64% median survival probability in patients without PH; these post-transplant patients without PH likely exhibit a 14-year median survival time comparative to the post-transplant national average (16). Juxtaposed against PH-LHD who had 7.5-year median survival, the impact of PH persistence on survival within this cohort appears to be strongly related to mPAP and PAWP at lower pulmonary pressures highlighting the impact of milder forms of PH (figure 4). The presence of post-transplant PH, specifically IpcPH, is likely due to a combination of factors such as cardiac donor characteristics, maladaptation in pulmonary vasculature not evident by RHC, hidden chronic transplant rejection related changes, or possibly consequences of donor-recipient pulmonary artery anastomoses mismatch secondary to surgery. Further studies are needed to investigate mechanisms impacting reduced survival in post-OHT patients with persistent PH.

Our study has several limitations. First, the retrospective nature of the study limits causal inference on the relationship between RVD and PH persistence given heterogeneity of treatment received in VAD and OHT patients and the variable number of repeated RHCs per treatment group. Cautious interpretation is required when comparing treatment groups to one another (VAD-DT, VAD-BTT, and OHT-alone) in terms of persistence of PH and related hemodynamic data given each group inherently possess its own selection bias, particularly survival bias in VAD-DT. Second, lack of comprehensive medication data, adjunctive mechanical therapies (intra-aortic balloon pump and RV assist device) are potential confounders which could under-or overestimate the PH prevalence and its impact on RVD. Third, missing RHC data was present in the study, largely at post-VAD RHC; repeated RHC post-VAD is not routine and missing randomly in a subset of VAD-BTT without clear indication, whereas VAD-DT patients had increased mortality in post-implantation period limiting further evaluation. Therefore, careful interpretation of the conclusions is prudent and post-VAD RHC surveillance requires further study. Fourth, limited serial hemodynamic data > 1 year after final intervention limited extended evaluation which could account for the time-varying effect of pulmonary pressures.

The strengths of our study include the longitudinal evaluation of RHC-based hemodynamics, impact of PH persistence based on PH-subtype and intervention received (VAD, OHT, or both), and the impact of PH persistence on survival. Our study is able to demonstrate the residual derangements of RV function and persistence of PH after each intervention and reduced survival in patients with persistent PH 1-year after final intervention. Further investigation is needed to confirm our findings in other similar cohorts and to evaluate whether hemodynamic optimization in VAD-assisted patients may reduce RVD and subsequently improve patient outcomes.

## Data Availability

All non-identifiable, non-personal health information data produced in the present study are available upon reasonable request to the authors.

## Acknowledgements

We would like to thank the Center for Translational Sciences Institute at the University of Pittsburgh for statistical support in the preparation of the manuscript which was supported in part by a grant from the National Institutes of Health (UL1-TR-001857).

## List of Abbreviations

PH: Pulmonary Hypertension
Cpc-PH: Combined Post-Capillary Pulmonary Hypertension
HF: Heart Failure
IpcPH: Isolated Post-Capillary Pulmonary Hypertension
LV: Left Ventricle
OHT: Orthotopic Heart Transplantation
PAH: Pulmonary Arterial Hypertension (Group 1 pulmonary hypertension)
PH-LHD: Pulmonary Hypertension in the setting of Left Heart Disease (Group 2 PH)
RV: Right Ventricle

