## Supplemental Materials for "Persistence of Pulmonary Hypertension in Patients undergoing Ventricular Assist Devices and Orthotopic Heart Transplantation"

**Arun Rajaratnam et al.**

### Supplemental Materials

### Supplemental Methods:

#### Evaluation of Invasive Hemodynamics

As detailed in the main text Methods section, right heart catheterization (RHC) data was used to elucidate proportion of patients with pulmonary hypertension (PH) and their respective subtypes in the patient cohorts of this study. These data are described in the Results section, Figures 1A-C and Supplemental Figures 1A-1C. For analyses of data in Figure 1, as described in the main text, we used the definition of PH as the total number of patients with PH burden based the presence of PH in the setting of HF (PH-LHD) or Pulmonary Arterial Hypertension (PAH) but not both (i.e diagnoses are mutually exclusive). PH-LHD was defined as mPAP>20 mmHg at rest in the supine position and PAWP ≥15mmHg. PAH was defined as mPAP > 20mmHg & PAWP < 15mmHg & PVR≥3 Woods Unit (WU). Presence of PH was evaluated in each treatment group in patients with available RHC and at each RHC period. The generalized linear model using the generalized estimating equation was used to evaluate longitudinal changes in the proportion of PH during the course of the study.

For analyses of data in Supplemental Figures 1A-1C we used stricter criteria to define subtypes of PH as noted in the methods section: PH-LHD (Group II PH) was further sub-categorized into Isolated post-capillary PH (IpcPH) defined as mPAP > 20 mmHg, PAWP ≥15mmHg and PVR <3 WU, and Combined post-capillary PH (CpcPH) defined as mPAP > 20mmHg, PAWP≥15mmHg and PVR>3 WU. Isolated elevated mPAP was defined as mPAP≥20 mmHg, PAWP < 15mmHg and PVR < 3 WU, and normal hemodynamics (i.e., no PH) defined as mPAP < 20mmg, PAWP < 15mmHg and PVR < 3 WU. This meant that PVR was necessary to include in these analyses, and therefore, only patients with available mPAP, PAWP, and PVR were included. As a result of this stricter definition, there were minor differences in data sample size between Figure 1 and Supplemental Figure 1. At pre-intervention RHC within VAD-BTT there was a difference of 16 patients, within VAD-DT 21 patients, and within OHT-alone this difference was 10 patients.

**Supplemental Data:**

**Supplemental Table 1: Comorbidities**

| | VAD-BTT N=111 | VAD-DT N = 164 | OHT-alone N= 138 | P value |
|---|---|---|---|---|
| Cardiac Arrythmia | 78% (39/50) | 68% (66/97) | 80% (45/56) | 0.19 |
| Cardiac Valvular Disease | 4% (2/50) | 11% (11/97) | 18% (10/56) | 0.08 |
| CKD | 12% (6/50) | 4% (4/97) | 4% (2/56) | 0.12 |
| COPD | 10% (5/50) | 12% (12/97) | 7% (4/56) | 0.59 |
| Diabetes Mellitus | 18% (9/50) | 30% (29/97) | 20% (11/56) | 0.18 |
| Hypertension | 48% (24/50) | 39% (38/97) | 30% (17/56) | 0.18 |
| Liver Disease | 4% (2/50) | 7% (7/97) | 4% (2/56) | 0.55 |
| PVD | 2% (1/50) | 11% (11/97) | 9% (5/51) | 0.15 |
| Obesity | 20% (10/50) | 11% (11/97) | 14% (8/56) | 0.36 |

Data are expressed as percent (%) and absolute value of patients with co-morbidity.
P-value < 0.05 was considered statistically significant using the Chi-square or Fisher's exact test.
Chronic Kidney Disease; COPD = Chronic Obstructive Pulmonary Disease; PVD = Peripheral Vascular Disease.

**Supplemental Figure 1A: Phenotypes of Pulmonary Hypertension in Advanced Heart Failure in VAD-BTT**

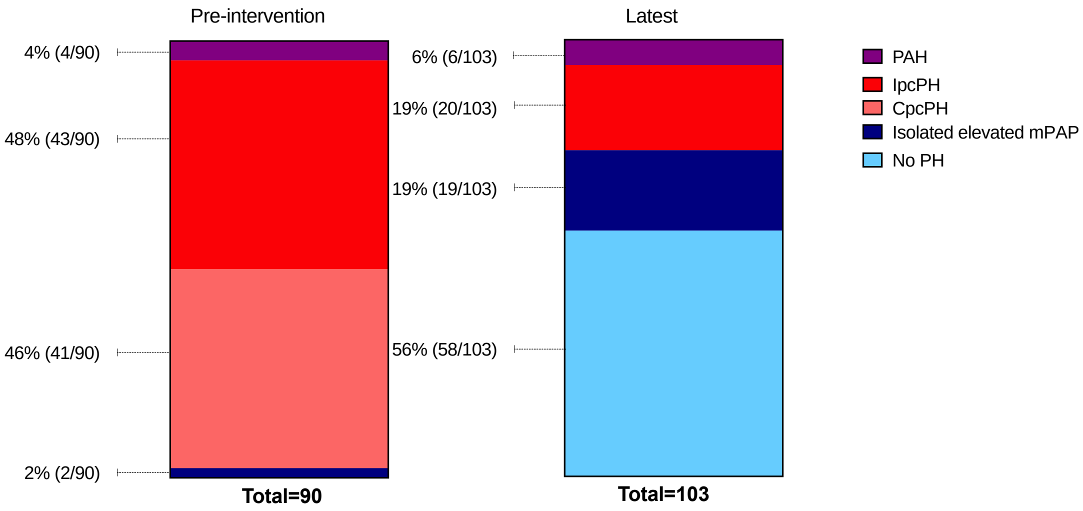


Proportion of patients expressed as absolute value with % exhibiting pulmonary arterial hypertension (P AH), Isolated Post-capillary pulmonary hypertenion (IpcPH), Combined post-capillary pulmonary hypertension (CpcPH), Isolated elevated mPAP elevation, and No Pulmonary Hypertension (No PH) based on treatment group prior to intervention (pre-intervention) and median 1 year after final intervention (latest RHC).

**Supplemental Figure 1B: Phenotypes of Pulmonary Hypertension in Advanced Heart Failure in VAD-DT**

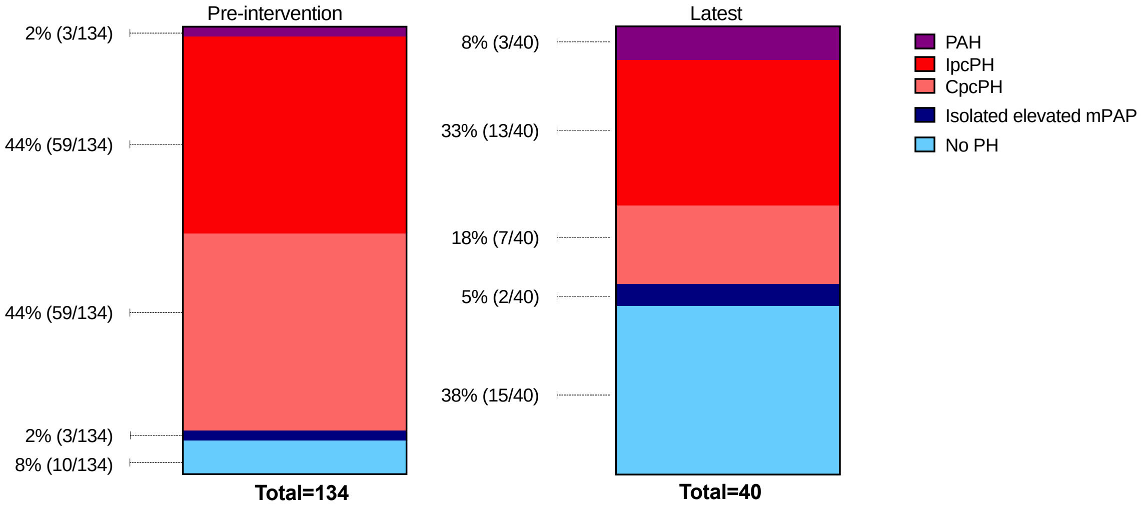


Proportion of patients expressed as absolute value with % exhibiting pulmonary arterial hypertension (P AH), Isolated Post-capillary pulmonary hypertenion (IpcPH), Combined post-capillary pulmonary hypertension (CpcPH), Isolated elevated mPAP elevation, and No Pulmonary Hypertension (No PH) based on treatment group prior to intervention (pre-intervention) and median 1 year after final intervention (latest RHC).

**Supplemental Figure 1C: Phenotypes of Pulmonary Hypertension in Advanced Heart Failure in OHT-alone**

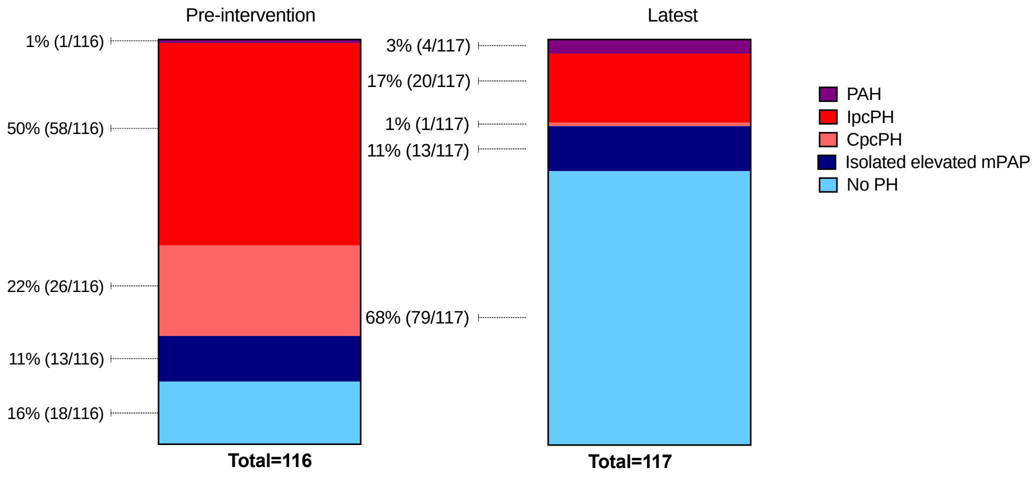


Proportion of patients expressed as absolute value with % exhibiting pulmonary arterial hypertension (P AH), Isolated Post-capillary pulmonary hypertenion (IpcPH), Combined post-capillary pulmonary hypertension (CpcPH), Isolated elevated mP AP elevation, and No Pulmonary hypertension (No PH) based on treatment group prior to intervention (pre-intervention) and median 1 year after final intervention (latest RHC).

**Supplemental Table 2: Right Ventricular Hemodynamics by Treatment Group at Latest-RHC**

| | VAD-BTT N=103 | VAD-DT N= 40 | OHT-alone N=118 | P value |
|---|---|---|---|---|
| RAP (mmHg) | 5 (3-7) | 10 (6)[a] | 4(3-7) | <0.01 |
| RASP (mmHg) | 7 (5-9) | 12 (6)[a] | 8 (4) | <0.01 |
| RADP (mmHg) | 8 (5-10) | 12 (8-17)[a] | 8 (6-10) | <0.01 |
| RVSP (mmHg) | 32 (8) | 40 (13)[a] | 32 (8) | <0.01 |
| RVDP (mmHg) | 1 (3) | 3 (0-7) [a] | 1 (2) | <0.01 |
| mRVP (mmHg) | 6 (4-8) | 10 (7-17)[a] | 6 (4-9) | <0.01 |
| RVSWI (mmHg*mL/ mm2) | 63 (66) | 131 (108)[a] | 64 (57) | <0.01 |

Values are expressed as mean (SD) or median (IQR). P-value was derived utilizing one-way ANOVA with Welch's test or the Kruskal-Wallis test. P-value < 0.05 was considered significant. Analyses for multiple comparisons were adjusted using the Bonferroni or Games-Howell method. [a] denotes VAD-DT vs VAD-BTT or OHT. RAP = right atrial pressure; RASP = right atrial systolic pressure, RADP = right atrial diastolic pressure, RVSP= right ventricular systolic pressure, RVDP= right ventricular diastolic pressure, mRVP = mean right ventricular pressure, RVSWI = right ventricular stroke volume index.

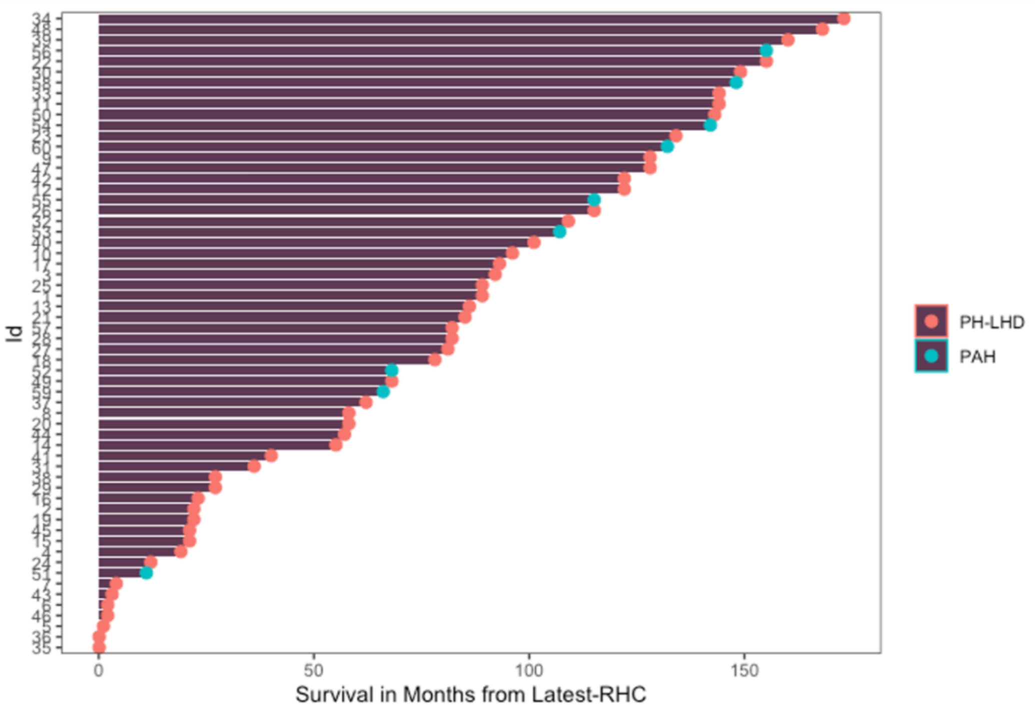


**Supplemental Figure 2: Swimmer's plot of Patient's with PH at post-cardiac transplant.** Swimmer's plot describes survival time in descending order of time (months) 1-year from cardiac transplant. Distribution of patients (Patient ID) with PAH subtype (green dot) shows even dispersion of mortality within the PH cohort, which rules out the possibility that PAH-associated early mortality could have had an impact on survival in patients with PH in the OHT cohort.

PAH = Pulmonary Arterial Hypertension; PH-LHD= PH secondary to left heart disease.

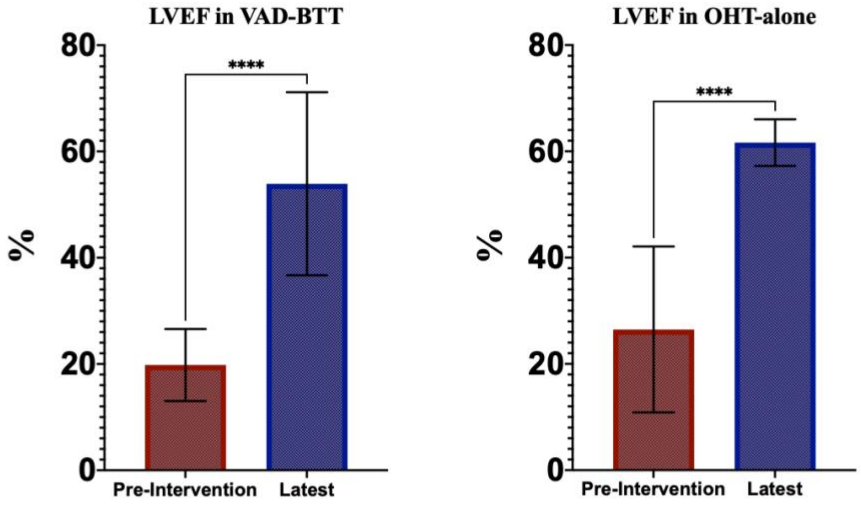


**Supplemental Figure 3.** Left Ventricular Ejection Fraction (LVEF) in VAD-BTT and OHT-alone groups show significant improvement by latest-RHC. LVEF reported as percentage (%) and panel arrows exhibit statistical signficance determined by Mann-Whitney U Test with *** denoting P-value of < 0.0001.

**Supplemental Table 3: Mortality statistics by Treatment Group between Pre-intervention RHC to Last-follow Up**

| | Alive | Death |
|---|---|---|
| **VAD as BTT** | 69% (77/111) | 31% (34/111) |
| **VAD as DT** | 27% (44/164) | 73% (120/164) |
| **OHT** | 65% (89/138) | 36% (49/138) |

Values are reported as percentage (%) with absolute values.

**Supplemental Figure 4: Definitions of Pulmonary Hypertension by Right Heart Catheterization**

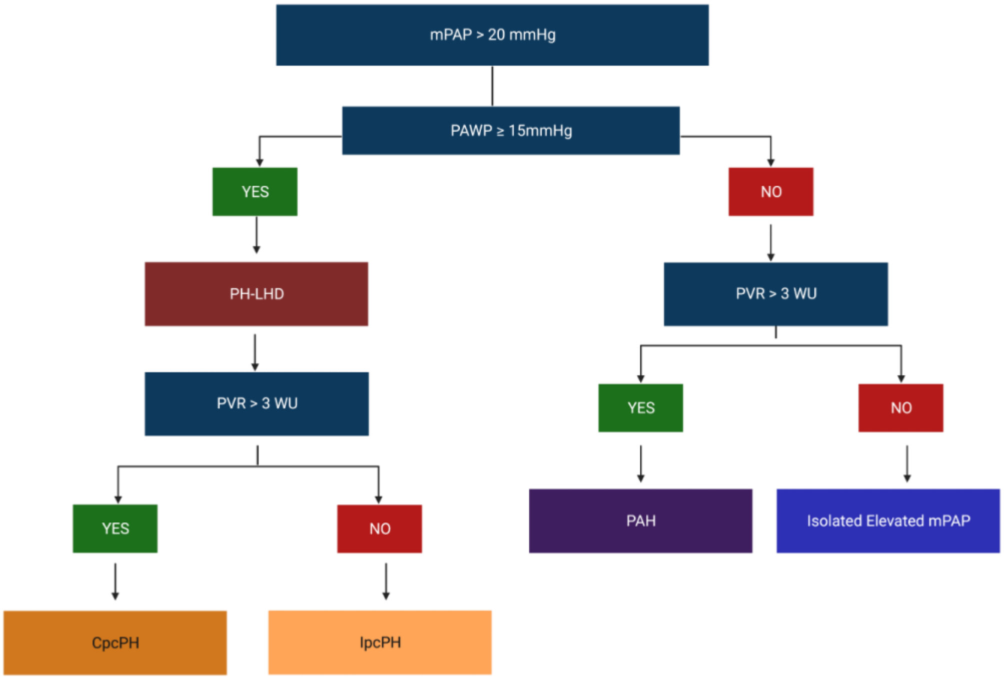


CpcPH : Combined Post-Capillary Pulmonary Hypertension, IpcPH : Isolated Post-Capillary Pulmonary Hypertension,PAH : Pulmonary Arterial Hypertension, PH-LHD = Pulmonary Hypertension in Left Heart Disease/Heart FailurePA :Pulmonary Artery; mPAP : mean PA pressure; PAWP :PA Wedge Pressure; PVR: Peripheral Vascular Resistance.
